# Effectiveness of Non-Pharmacological Interventions to Improve Cognition in Cancer: A Protocol for an Overview of Systematic Reviews

**DOI:** 10.1101/2024.04.02.24305172

**Authors:** Darren Haywood, Ashley M. Henneghan, Evan Dauer MclinPsych, Sherry Vasan, Oscar Y. Franco-Rocha, Helen Wilding, Nicolas H. Hart

## Abstract

Many cancer survivors, including those with a current or previous diagnosis of cancer, experience cancer-related cognitive impairments (CRCI). CRCI can impact their ability to think quickly, clearly, make decisions and perform daily actions. There have been a variety of non-pharmacological interventions developed and trialed with the aim of reducing CRCI or mitigating its impact. The aim of this work is to provide an overall picture of the effectiveness of non-pharmacological interventions to improve cognition in cancer survivors by providing an overview and synthesis of systematic reviews.

**Review Title:** Effectiveness of Non-Pharmacological Interventions to Improve Cognition in Cancer: An Overview of Systematic Reviews

## Background

A significant proportion of cancer survivors, defined as anyone who has had a cancer diagnosis, experience cognitive impairments that can impact their ability to think quickly, clearly, make decisions and perform daily actions [1-3]. This is commonly called cancer-related cognitive impairment (CRCI) [1]. It is largely believed that cancer, cancer treatments, and psychosocial factors may all contribute to the development and maintenance of CRCI [4]. Cognitive impairment is also generally related to a variety of negative physical and psychological outcomes across populations [5-10], and cancer survivors are at a heightened risk of experiencing these outcomes, such as psychopathology [11]. To reduce CRCI and minimise its impact, a wide variety of non-pharmacological interventions have been trialed across human and animal cancer survivor populations including exercise, cognitive behavioural therapy, cognitive training, dietary, and mindfulness interventions [4]. There have now been several systematic reviews published, both broader and narrower in scope, that have examined the effectiveness of various non-pharmacological interventions aimed at improving cognition in cancer survivors of various types. To provide an overall picture of the effectiveness of non-pharmacological interventions to improve cognition in cancer survivors an overview and synthesis of these systematic reviews is required. This overview may inform clinical guidelines and educational responses, as well as point toward the most promising interventions and uncover gaps in the current systematic review literature.

## Objective

To provide a comprehensive overview of systematic reviews examining the effectiveness of any non-pharmacological interventions aimed at improving cognition in cancer populations.

### Anticipated or Actual Start Date

12/03/24

### Anticipated Completion Date

15/08/24

### Named Contact

Dr Darren Haywood

### Organizational Affiliation of the Review

University of Technology Sydney

### Review Team Members and their Organizational Affiliations

#### PI

Name: Darren Haywood, PhD, BPsych(Hons), Dip. Fit

Postdoctoral Research Fellow (Cancer Survivorship)

Human Performance Research Centre

#### Co-PI

Name: Nicolas H. Hart, PhD AES CSCS ESSAF, Associate Professor

NHMRC Investigator Fellow (Cancer Survivorship)

Program Director (Clinical Exercise Physiology)

Human Performance Research Centre

University of Technology Sydney (UTS)

#### Investigator

Name: Ashley M. Henneghan, PhD, RN, Associate Professor

School of Nursing

University of Texas at Austin

#### Investigator

Name: Evan Dauer, MClinPsych, BSc(Hons)

Research Fellow

Human Performance Research Centre

#### Investigator

Name: Sherry Vasan, PhD, BSc(Hons)

Research Fellow

Human Performance Research Centre

#### Investigator

Name: Oscar Y. Franco-Rocha, M.Ed., RN, PhD Student

School of Nursing

University of Texas at Austin

### Funding sources/sponsors

There is no financial support for this research.

### Investigator

Name: Helen Wilding

Senior Research Librarian

St. Vincent’s Hospital Melbourne

### Funding sources/sponsors

There is no financial support for this research.

## Conflicts of interest

None.

### Collaborators

None.

### Review Question

What is the effectiveness of non-pharmacological interventions for improving cognition in cancer populations?

### Searches

Medline ALL (Ovid), Embase (Ovid), Emcare (Ovid), APA PsycINFO (Ovid), CINAHL (EBSCOhost), Cochrane Library (Wiley). Search date 12/03/24.

No age restrictions. To identify additional potentially relevant studies the reference list of included systematic reviews will be hand-searched.

Exact searches are presented below for each database.

### Ovid MEDLINE(R) ALL 1946 to March 12, 2024

1. exp Neoplasms/ or Cancer Survivors/ or Cancer Care Facilities/ or Oncology Service, Hospital/ or exp Medical Oncology/ or Oncology Nursing/ or Cancer Pain/
2. (cancer* or carcinoma* or glioblastoma* or leuk?emia* or lymphoma* or malignan* or neoplasm* or tumo?r or tumo?rs or oncolog*).ti,ab,kf.
3. 1 or 2
4. cognition/ or awareness/ or cognitive reserve/ or comprehension/ or metacognition/ or processing speed/
5. cognition disorders/ or cognitive dysfunction/ or chemotherapy-related cognitive impairment/ or postoperative cognitive complications/
6. attention/ or executive function/
7. memory/ or memory, episodic/ or memory, long-term/ or memory, short-term/ or mental recall/
8. Mental Processes/ or Neurobehavioral Manifestations/ or exp Neuropsychological Tests/ or Problem Solving/
9. thinking/ or concept formation/ or judgment/
10. (chemobrain or chemo-brain or chemofog or chemo-fog or cognit* or concept formation or executive function* or metacognit* or memory or mental process*).ti,ab,kf.
11. (neurobehavioral manifest* or neurocognit* or neuropsychological test* or processing speed).ti,ab,kf.
12. 4 or 5 or 6 or 7 or 8 or 9 or 10 or 11
13. meta-analysis/ or “systematic review”/ or Systematic Reviews as Topic/
14. ((system* or literature) adj2 (review* or search)).ti,ab,kf.
15. (cochrane review or meta-analysis or meta-synthesis or narrative review* or quantitative review* or systematic overview* or prisma or medline or pubmed or embase).ti,ab,kf.
16. (meta analysis or “systematic review”).pt.
17. 13 or 14 or 15 or 16
18. 3 and 12 and 17
19. (case reports or comment or editorial or letter or news).pt.
20. 18 not 19

### Embase 1974 to 2024 March 12 (Ovid)

1. exp *neoplasm/ or *cancer survivor/ or *cancer patient/ or exp *cancer survival/ or exp *cancer therapy/ or *cancer center/ or exp *oncology/ or *oncology ward/
2. (cancer* or carcinoma* or glioblastoma* or leuk?emia* or lymphoma* or malignan* or neoplasm* or tumo?r or tumo?rs or oncolog*).ti,ab,kf.
3. 3., 1 or 2
4. *chemotherapy-related cognitive impairment/ or *cognitive defect/ or *mild cognitive impairment/ or *postoperative cognitive dysfunction/
5. *cognition/ or *attention/ or *cognitive reserve/ or *executive function/ or *memory/ or *metacognition/ or *processing speed/ or *thinking/ or exp *neuropsychological assessment/
6. (chemobrain or chemo-brain or chemofog or chemo-fog or cognit* or concept formation or executive function* or metacognit* or memory or mental process*).ti,ab,kf.
7. (neurobehavioral manifest* or neurocognit* or neuropsychological test* or processing speed).ti,ab,kf.
8. 4 or 5 or 6 or 7
9. “systematic review”/ or “systematic review (topic)”/ or meta analysis/ or “meta analysis (topic)”/
10. ((system* or literature) adj2 (review* or search)).ti,ab,kf.
11. (cochrane review or meta-analysis or meta-synthesis or narrative review* or quantitative review* or systematic overview* or prisma or medline or pubmed or embase).ti,ab,kf.
12. 9 or 10 or 11
13. 3 and 8 and 12
14. (book or chapter or conference or conference abstract or conference paper or “conference review” or editorial or letter or note).pt.
15. case report.ti.
16. 14 or 15
17. 13 not 16

### Ovid Emcare 1995 to 2024 Week 10

1. exp *neoplasm/ or *cancer survivor/ or *cancer patient/ or exp *cancer survival/ or exp *cancer therapy/ or *cancer center/ or exp *oncology/ or *oncology ward/
2. (cancer* or carcinoma* or glioblastoma* or leuk?emia* or lymphoma* or malignan* or neoplasm* or tumo?r or tumo?rs or oncolog*).ti,ab,kf.
3. 1 or 2
4. *chemotherapy-related cognitive impairment/ or *cognitive defect/ or *mild cognitive impairment/ or *postoperative cognitive dysfunction/
5. *cognition/ or *attention/ or *cognitive reserve/ or *executive function/ or *memory/ or *metacognition/ or *processing speed/ or *thinking/ or exp neuropsychological test/
6. (chemobrain or chemo-brain or chemofog or chemo-fog or cognit* or concept formation or executive function* or metacognit* or memory or mental process*).ti,ab,kf.
7. (neurobehavioral manifest* or neurocognit* or neuropsychological test* or processing speed).ti,ab,kf.
8. 4 or 5 or 6 or 7
9. “systematic review”/ or “systematic review (topic)”/ or meta analysis/ or “meta analysis (topic)”/
10. ((system* or literature) adj2 (review* or search)).ti,ab,kf.
11. (cochrane review or meta-analysis or meta-synthesis or narrative review* or quantitative review* or systematic overview* or prisma or medline or pubmed or embase).ti,ab,kf.
12. 9 or 10 or 11
13. 3 and 8 and 12
14. (book or chapter or conference or conference abstract or conference paper or “conference review” or editorial or letter or note).pt.
15. case report.ti.
16. 14 or 15
17. 13 not 16

### APA PsycInfo 1806 to March Week 2 2024 (Ovid)

1. exp neoplasms/ or oncology/
2. (cancer* or carcinoma* or glioblastoma* or leuk?emia* or lymphoma* or malignan* or neoplasm* or tumo?r or tumo?rs or oncolog*).ti,ab.
3. 1 or 2
4. cognitive processes/ or exp awareness/ or exp cognition/ or exp cognitive ability/ or cognitive processing speed/ or cognitive reserve/ or exp cognitive strategies/ or concentration/ or exp concept formation/ or exp executive function/ or exp judgment/ or metacognition/ or exp problem solving/ or exp thinking/ or exp cognitive assessment/
5. cognitive impairment/ or mild cognitive impairment/ or neurocognitive disorders/ or memory/
6. (chemobrain or chemo-brain or chemofog or chemo-fog or cognit* or concept formation or executive function* or metacognit* or memory or mental process*).ti,ab.
7. (neurobehavioral manifest* or neurocognit* or neuropsychological test* or processing speed).ti,ab.
8. 4 or 5 or 6 or 7
9. “systematic review”/ or “literature review”/ or meta analysis/
10. ((system* or literature) adj2 (review* or search)).ti,ab.
11. (cochrane review or meta-analysis or meta-synthesis or narrative review* or quantitative review* or systematic overview* or prisma or medline or pubmed or embase).ti,ab.
12. 9 or 10 or 11
13. 3 and 8 and 12

### CINAHL (EBSCOhost)

S1. (MH “Cancer Survivors”) OR (MH “Cancer Patients”) OR (MH “Cancer Fatigue”) OR (MH “Cancer Pain”) OR (MH “Chemotherapy, Cancer+”) OR (MH “Rehabilitation, Cancer”)
S2. (MH “Neoplasms+”) OR (MH “Oncology+”) OR (MH “Oncology Nursing+”)
S3. cancer* OR carcinoma* OR glioblastoma* OR leuk#emia* OR lymphoma* OR malignan* OR neoplasm* OR tumo#r OR tumo#rs OR oncolog*
S4. S1 OR S2 OR S3
S5. (MH “Cognition Disorders+”) OR (MH “Chemotherapy-Related Cognitive Impairment”) OR (MH “Mild Cognitive Impairment”)
S6. (MH “Cognition”) OR (MH “Processing Speed”) OR (MH “Executive Function”) OR (MH “Mental Processes”) OR (MH “Thinking”)
S7. (MH “Problem Solving”) OR (MH “Memory”) OR (MH “Neuropsychological Tests+”)
S8. chemobrain OR chemo-brain OR chemofog OR chemo-fog OR cognit* OR “concept formation” OR “executive function*” OR metacognit* OR memory OR “mental process*”
S9. “neurobehavioral manifest*” OR neurocognit* OR “neuropsychological test*” OR “processing speed”
S10. S5 OR S6 OR S7 OR S8 OR S9
S11. (MH “Literature Review+”) OR (MH “Systematic Review”) OR (MH “Scoping Review”) OR (MH “Meta Analysis”)
S12. (system* OR literature) N2 (review* OR search)
S13. “cochrane review” OR meta-analysis OR meta-synthesis OR “narrative review*” OR “quantitative review*” OR “systematic overview*” OR prisma OR medline OR pubmed OR embase
S14. S11 OR S12 OR S13
S15. S4 AND S10 AND S14

### Cochrane Library (Wiley)

1. [mh Neoplasms] OR [mh ^”Cancer Survivors”] OR [mh ^”Cancer Care Facilities”] OR [mh ^”Oncology Service, Hospital”] OR [mh “Medical Oncology”] OR [mh ^”Oncology Nursing”] OR [mh ^”Cancer Pain”]
2. cancer*:ti,ab OR carcinoma*:ti,ab OR glioblastoma*:ti,ab OR leuk?emia*:ti,ab OR lymphoma*:ti,ab OR malignan*:ti,ab OR neoplasm*:ti,ab OR tumo?r:ti,ab OR tumo?rs:ti,ab OR oncolog*:ti,ab
3. #1 OR #2
4. [mh ^cognition] OR [mh ^awareness] OR [mh ^”cognitive reserve”] OR [mh ^comprehension] OR [mh ^metacognition] OR [mh ^”processing speed”]
5. [mh ^”cognition disorders”] OR [mh ^”cognitive dysfunction”] OR [mh ^”chemotherapy-related cognitive impairment”] OR [mh ^”postoperative cognitive complications”]
6. [mh ^attention] OR [mh ^”executive function”]
7. [mh ^memory] OR [mh ^”memory, episodic”] OR [mh ^”memory, long-term”] OR [mh ^”memory, short-term”] OR [mh ^”mental recall”]
8. [mh ^”Mental Processes”] OR [mh ^”Neurobehavioral Manifestations”] OR [mh “Neuropsychological Tests”] OR [mh ^”Problem Solving”]
9. [mh ^thinking] OR [mh ^”concept formation”] OR [mh ^judgment]
10. chemobrain:ti,ab OR chemo-brain:ti,ab OR chemofog:ti,ab OR chemo-fog:ti,ab OR cognit*:ti,ab OR “concept formation”:ti,ab OR (“executive” NEXT function*):ti,ab OR metacognit*:ti,ab OR memory:ti,ab OR (“mental” NEXT process*):ti,ab
11. (“neurobehavioral” NEXT manifest*):ti,ab OR neurocognit*:ti,ab OR (“neuropsychological” NEXT test*):ti,ab OR “processing speed”:ti,ab
12. #4 OR #5 OR #6 OR #7 OR #8 OR #9 OR #10 OR #11
13. (system*:ti,ab OR literature:ti,ab) NEAR/2 (review*:ti,ab OR search:ti,ab)
14. “cochrane review”:ti,ab OR meta-analysis:ti,ab OR meta-synthesis:ti,ab OR (“narrative” NEXT review*):ti,ab OR (“quantitative” NEXT review*):ti,ab OR (“systematic” NEXT overview*):ti,ab OR prisma:ti,ab OR medline:ti,ab OR pubmed:ti,ab OR embase:ti,ab
15. MeSH descriptor: [Systematic Review] explode all trees
16. #13 OR #14 OR #15
17. #3 AND #12 AND #16

### PICO

#### Participants/Population

Populations who have had a cancer diagnosis, either current or previous. Any stage of the cancer journey. All types of cancer. All types of treatments. All ages. All genders/sexes. Humans or animals.

#### Intervention(s), Exposure(s)

Non-pharmacological interventions aimed to improve cognition.

#### Comparator(s)/Control

Must include a within or between group comparison. All comparisons are included (i.e., to a control group, pre-post, etc).

#### Outcome

Cognitive functioning (subjective and/or objective)

### Condition or Domain Being Studied

Cognition, Cancer, non-pharmacological interventions.

### Types of Study to be Included

Systematic reviews reporting the effectiveness of non-pharmacological interventions to improve cognition in cancer populations.

‘Systematic review’ was defined using the PRISMA-P statement definition of a SR: (A) had an explicit aim; (B) used a systematic search strategy and selection of studies; and (C) systematically synthesized data using narrative synthesis and/or meta-analysis. Unpublished work, abstracts, letters, and conference proceedings were not included.

### Exclusion Criteria

Systematic reviews only including studies with pharmacological interventions (including supplements available over the counter) aimed at improving cognition.

Systematic reviews that do not report on subjective or objective measures of cognition.

Non-systematic reviews.

### Inclusion Criteria

Systematic reviews examining the effectiveness of any non-pharmacological interventions aimed at improving cognition in cancer populations.

Human or animal systematic reviews.

Have been diagnosed with any type of cancer (current or previous), any stage of cancer encompassing the entire cancer trajectory.

All ages of participants included.

Systematic reviews that include Subjective and/or objective measures of cognition

Systematic reviews that include any comparisons to establish intervention effectiveness (i.e., pre-post, control group, etc.)

Quality of Life (QoL) and other wellbeing systematic reviews in which at least 50% of included primary studies have a cognitive outcome measure.

Multiple population reviews where cancer is separately analyzed/synthesized and at least 50% of included primary cancer studies have a cognitive outcome measure.

### Data Extraction (Selection and Coding)

Study selection and data extraction will follow the Overview of Reviews in Cochrane Handbook for Systematic Reviews of Interventions guidelines [12] and will be reported per PRISMA 2020 statement [13]. The results will be imported to Covidence to remove duplicates and screening. Two reviewers will independently screen the studies in two stages: (1) title/abstract and (2) full texts. Disagreement will be resolved by a third reviewer.

Study characteristics to be extracted will include publication year, title, authors, journal, review type, country, interventions included, length of intervention(s), human or animal, cancer types, cancer treatments, cancer stage(s), comparison type(s), cognitive measures included, QoL and patient reported outcome measure(s) (PROMs) included, effectiveness in improving cognition, effectiveness in improving QoL and patient reported outcomes (PROs).

### Risk of Bias (Quality) Assessment

Two investigators will independently conduct a systematic review quality assessment using the Assessment of Multiple SRs (AMSTAR) 2 Checklist, a 16-question tool that evaluates each item as “yes” or “no” and yields a final overall rating of “high,” “moderate,” “low,” or “critically low” quality. Study quality disagreements were managed by consensus between authors and resolved by discussion with a third author if necessary.

### Data Synthesis and Analysis

Narrative Synthesis.

Corrected Covered Area (CCA) will be calculated to determine the extent to which primary studies overlap in the included SRs.

### Analysis of Subgroups or Subsets

Types of non-pharmacological interventions included within the systematic reviews will be grouped (i.e., exercise, cognitive training, cognitive behavioural therapy, etc). The results will be narratively synthesised both within and between these groups.

### Strategy for Data Synthesis

Data synthesis will take place using a narrative synthesis approach. No additional re-analysis of outcome data was conducted. Synthesis will be conducted both across intervention types and within intervention types.

### Type and Method of Review

Overview of Systematic Reviews “umbrella review”

### Health Area of the Review

Cancer and oncology

### Language

English

### Country

Australia; United States and America.

### Ethical Considerations

Not required.

### Dissemination Plans

The results will be published in a peer-reviewed journal and presented at academic conferences.

## Data Availability

All data produced in the present work will be contained in the manuscript

